# Value-based pricing of a COVID-19 vaccine

**DOI:** 10.1101/2021.03.06.21253035

**Authors:** Afschin Gandjour

**Affiliations:** Frankfurt School of Finance & Management, Frankfurt, Germany

**Keywords:** value-based price, COVID-19 vaccine

## Abstract

**Aim:** The purpose of this study is to determine the value-based price of a COVID-19 vaccine from a societal perspective in Germany.

**Methods:** A decision model was constructed using, e.g., information on age-specific fatality rates, intensive care unit (ICU) costs and outcomes, and herd protection threshold. Three strategies were analysed: vaccination (with 95% and 50% efficacy), a mitigation strategy, and no intervention. The base-case time horizon was 5 years. The value of a vaccine included savings from avoiding COVID-19 mitigation measures and health benefits from avoiding COVID-19 related mortality. The value of an additional life year was borrowed from new, innovative oncological drugs, as cancer reflects a condition with a similar morbidity and mortality burden in the general population in the short term as COVID-19.

**Results:** A vaccine with a 95% efficacy dominates the mitigation strategy strictly. The value-based price (€1494) is thus determined by the comparison between vaccination and no intervention. This price is particularly sensitive to the probability of ICU admission and the herd protection threshold. In contrast, the value of a vaccine with 50% efficacy is more ambiguous.

**Conclusion:** This study yields a value-based price for a COVID-19 vaccine with 95% efficacy, which is more than 50 times greater than the purchasing price.

## Introduction

In view of the second wave of severe acute respiratory syndrome coronavirus 2 (SARS-CoV-2) pandemic, the German federal government and federal states are pursuing a COVID-19 mitigation strategy (Bundesregierung 2020). It includes measures such as partial shutdown of businesses, social distancing, contact tracking, testing, public mask wearing, and quarantine orders (Bundesregierung 2020). An important goal of this strategy is to control COVID-19 outbreaks or postpone them (‘flatten the curve’) and thus avoid over-stretching the intensive care unit (ICU) capacity at peak demand (cf. Bundesregierung 2020).

The pharmaceutical companies Pfizer/Biontech and Moderna have independently shown through randomized double-blind placebo-controlled clinical trials (n = 43,548 and n = 30,351, respectively) that mRNA-based vaccine candidates against SARS-CoV-2 are about 95% effective in preventing symptomatic COVID-19 in participants who showed no evidence of prior infection with SARS-CoV-2 (Polack 2020, FDA 2020). The European Union approved the Biontech-Pfizer and Moderna vaccines for use across the 27 Member States on December 21, 2020 and January 6, 2021, respectively. However, the process of developing, approving, and distributing these and other vaccines may take until the end of 2021 in Germany (BBC 2020).

The estimated cost of immunizing one person is €26 based on €8.9 billion total acquisition expenditures borne by the German government (DW 2020) to vaccinate 336 million people (. Given the pressing public health needs, manufacturers do not seek maximum returns (cf. COVAX Facility 2020) and therefore, the prices are cost-based rather than value-based (Towse 2020). Value-based pricing (VBP) sets the prices of medical technologies based on various factors, including the health benefits they provide. According to a narrow definition of VBP, an explicit willingness-to-pay threshold is used to determine the price (Bouvy 2013). The purpose of this study is to determine the value-based price of a COVID-19 vaccine for use in the German general population from a societal perspective. The difference between the cost-based price and the value-based price helps in quantifying the social contribution of the manufacturers.

## Methods

### Pricing framework

In a VBP framework that relies on an economic evaluation, the maximum acceptable price of a new vaccine is determined by equating the incremental cost-effectiveness ratio (ICER) of the new vaccine compared with a less effective treatment to the cost-effectiveness threshold λ:

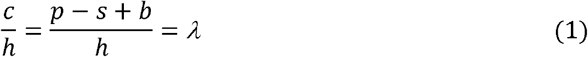

where *c* is incremental costs; *h* denotes incremental net health benefit including harm from side effects; *P* is the maximum acceptable additional price of the new vaccine net of the costs of the comparator as well as costs of vaccination, subsidies, establishing vaccination centres, transportation, and managing side effects; *s* denotes savings from avoiding COVID-19-related morbidity and COVID-19 mitigation measures; and *b* is the cost resulting from avoidance of premature death. Given the consideration of subsidies, the value-based price is adjusted downwards for public push for research and development (R&D) and manufacturing funding (cf. Towse 2020). Of note, the real-world costs of purchasing the vaccines and the costs of scientific research failures are not included in the calculation of the value-based price because they matter only for cost-plus pricing and not for VBP.

Rearranging Eq. (1) yields the maximum acceptable price of a new vaccine (Gandjour 2011):

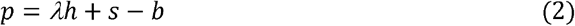

Note that *p* is calculated over the model’s time horizon. To derive the cost per dose, *p* needs to be divided by the number of doses over the time horizon.

In the following, I will call a VBP rule that applies an absolute cost-effectiveness threshold ‘absolute’ rule. In contrast, a proportional rule for VBP sets that costs induced by a vaccine should (only) increase in proportion to its incremental health benefits (Gandjour 2011). This rule was recently validated (Gandjour 2020). It implies a constant trade-off between costs and health benefits as shown in the following:

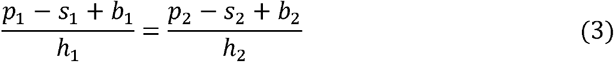

If the comparator of a COVID-19 vaccine are mitigation and ‘no intervention’, then subscript 1 denotes the comparison between the mitigation strategy and ‘no intervention’ and subscript 2 refers to the comparison between COVID-19 vaccine and the mitigation strategy. Importantly, comparators need to be non-dominated.

Solving for *p*_2_ yields the value-based price of a COVID-19 vaccine:

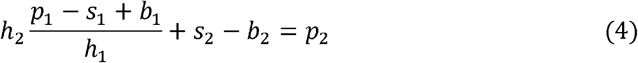

In this study, both rules are applied to derive the value-based price of a COVID-19 vaccine.

### Comparators

To apply the proportional rule, the study uses two comparators of a COVID-19 vaccine, ‘no intervention’ and the current practice in Germany, which is the mitigation strategy including a partial lockdown/shutdown.

In the short term, a vaccination strategy must be regarded as an add-on to a mitigation strategy because vaccination of a large part of the population cannot be achieved immediately. However, in the mid- to long-term, vaccination avoids the costs of mitigation strategy, which is the contribution of the lockdown/shutdown to the total economic burden of the SARS-CoV-2 pandemic. In addition, vaccination avoids the deaths associated with mitigation strategy, which is not able to suppress the pandemic. Nevertheless, a vaccine with only 50% efficacy may still require imposing lockdown measures even in the long-term (Reuters 2020). In a sensitivity analysis, the costs and benefits of the latter were included.

From the perspective of static efficiency, the GDP drop associated with the lockdown/shutdown till today can be considered sunk, while from the perspective of dynamic efficiency, which sets incentives for innovations (e.g., for vaccines in future pandemics), it is still relevant. As in future pandemics vaccine development and distribution is likely to occur only in conjunction with a mitigation strategy, considering the full mitigation cost avoids introducing excessive incentives for innovation. Therefore, a dynamic efficiency perspective was considered in the base case.

As the current mitigation strategy is not economically sustainable in the long run, ‘no intervention’ is chosen as a more realistic long-term comparator. ‘No intervention’ lifts the lockdown/shutdown and results in herd immunity through natural infection. Considering that an uncontrolled pandemic in Germany would require a peak capacity of several hundred thousand ICU beds (Khailaie 2020), overburdening of ICUs was assumed, leading to voluntary restrictions on economic activities. Many German commentators have hypothesized such a response following the pictures from the main hospital in the Italian city of Bergamo in March 2020.

### Decision model

A decision model was constructed based on a previously developed and validated model (Gandjour 2020a). The latter model determines the gain in life years of a strategy that is successful in ‘squashing the curve’ compared to the situation before the pandemic. It is based a life-table model that summarizes the age-specific mortality impact of the SARS-CoV-2 pandemic. The base-case calculation relies on an independence assumption, implying that individuals not dying from COVID 19 have the same probability of death as all individuals before the rise of the pandemic. Given that patients who die from COVID 19 tend to have more comorbidities (Wu 2020), I assumed a harvesting effect in a sensitivity analysis. This approach presumes that those who die from COVID-19 are sicker and would have died any-way. In this scenario, I assumed for age groups with excess mortality associated with COVID-19 (the difference between observed and pre-pandemic mortality rates) that except for COVID-19, there are no other causes of death in the forthcoming 12 months. To account for the age distribution of the population, the model weighs age-specific life-expectancy changes by age-specific population sizes. As the strategy chosen by the German government tends toward mitigation than suppression, I adjusted the number of life years gained from ‘squashing the curve’ for the expected number of pandemic waves and the resulting death tolls under mitigation.

The time horizon (5 years in the base case) was set based on the expected duration of immunity. The transmission dynamics of SARS-CoV-2 are comparable to those of influenza (van Damme 2020), which typically causes epidemics in temperate climates every year during winter. In the absence of a vaccine, future SARS-CoV-2 pandemic waves were therefore assumed to peak in winter and return yearly.

### Vaccine efficacy

Vaccine efficacy can be defined based on the attack rate (the proportion of individuals infected in the specific risk group over a nominated period) or the frequency of only severe cases (Préziosi 2003). The herd immunity threshold was calculated based on an inversely proportional relationship with vaccine efficacy (in terms of attack rate) (Chowell 2009):

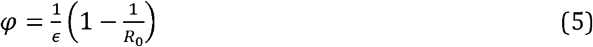

where *φ* refers to the herd immunity threshold, *∊* is vaccine efficacy, and *R*_O_ is the basic reproduction number of a disease.

### Cost calculation

The study took a societal viewpoint, by including both direct medical costs and indirect/productivity costs. The savings in health care expenditures by avoiding the spread of SARS-CoV-2 in the population under no intervention were not added to the GDP estimate, because the savings were assumed to be offset by higher health expenditures for elective procedures and emergency and physician visits for unrelated medical conditions. That is, in case of a natural spread, providers and patients were assumed to reduce utilization of elective procedures as well as emergency and physician visits for unrelated medical conditions.

Conversely, productivity gains from avoiding sickness by mitigation and vaccination were added to the GDP estimates. They were calculated by multiplying the proportion of symptomatic patients with the duration of sickness and the daily productivity loss. For the latter calculation a few simplifying assumptions were made. First, all undiagnosed but symptomatic infections were assumed to be mild. This assumption is indirectly supported by official data on excess mortality in Germany (Federal Office of Statistics 2020) showing that the peak in excess mortality in the first half of April and the rising mortality in November both coincided with surges in COVID-19 deaths (thus essentially ruling out deaths due to undiagnosed COVID-19 cases). Second, the maximum number of quarantined contact persons per diagnosed reached the maximum in August 2020 and cannot increase further due to labor and technological constraints in local public health departments. Third, infected and quarantined individuals are representative of the general population (which is a plausible assumption because 98% of the German population has not been infected thus far (Robert Koch Institut 2020)).

## Data

### Economic data

According to the European Economic Forecast by the European Commission in November 2020, GDP of Germany is set to contract by 5.5% in 2020. Second wave of infections is expected to dampen the rebound to 3.5% next year. Assuming there is no permanent damage to productive capacity, Germany’s economy is projected to continue to grow above potential in 2022 at 2.5% and complete its recovery to the pre-crisis levels. As the 2021 GDP growth projection was revised down to 3.5% from 5.3% in the forecast of July 2020, the impact of the second wave is calculated to be a 1.8% contraction of GDP. This percentage was also applied to potential future waves. According to the European Economic Forecast, the total volume of the government measures “to fight the COVID-19 pandemic and stabilise the economy (…) amounts to 4.7% of GDP in 2020 and 2.1% in 2021”. By subtracting the GDP contraction due to the second wave, I determined the GDP loss independent of the second wave.

However, the European Economic Forecast was conducted assuming the absence of a pandemic in the counterfactual scenario, without considering the voluntary restrictions such as social distancing in view of the rapid spread of the virus in the population (cf. Aum 2020). That is, individuals may take precautions even without the lockdown orders. Accounting for the latter would decrease the incremental cost of the lockdown/shutdown over no pandemic. In a sensitivity analysis I assumed the contribution of the lockdown/shutdown to the total loss of economic activities to be 10%, to account for the voluntary restrictions that may take place in the absence of a lockdown/shutdown. This estimate agrees with the one regarding the contribution of a shutdown to the loss of economic activities in Denmark, which was estimated to be 14% (=4%/29%) (Sheridan 2020).

To determine the productivity gains resulting from a vaccination or mitigation strategy, as compared to no intervention, I used the data sources reported in Table A1 of the Appendix.

The German federal government has been funding three vaccine developers with a total of 750 million euros. BioNTech from Mainz received 375 million euros and Curevac from Tübingen received 230 million euros through a special vaccine development program (Zeit 2020). In addition, the federal government is planning one billion euros for the construction and operation of the planned vaccination centers (RND 2020). Both types of costs were included in the analysis and related to one vaccinated individual. The present study assumes that vaccination mainly takes place in vaccination centers.

The number of acquired doses is 635 million (Tagesschau 2021), which would allow vaccination of 336 million people assuming up to 2 doses per individual. If reaching herd immunity based on 100% efficacy requires 70% of the population to be infected (Kwok 2020), 66% of doses will not be used. Nevertheless, due to the existing purchase agreements the value-based price also needs to include the unused doses. Effectively, the value-based price thus needs to be reduced by the proportion of unused vaccines.

All costs are presented in euros, year 2020 values.

### Clinical and epidemiological data

In agreement with other authors who foresaw a high probability of a second and third SARS-CoV-2 pandemic wave under no intervention strategy (e.g., Fakhruddin 2020, Pollock 2020), I assumed two remaining pandemic waves under no intervention in the base case.

To calculate the per capita gain in life years through mitigation, I applied the currently estimated COVID-19 infection fatality rate (IFR) of 0.75% (WHO 2020) to the previously developed model (Gandjour 2020a). To account for the death toll under mitigation I assumed in the base case that the number of daily COVID-19 deaths since December 11, 2020 (approximately 600) will remain constant until the end of March and then subsides. While it appears likely that a significantly higher number of deaths would not be tolerated because of concerns about overburdening of the ICU capacity, the number would be reduced by more than two thirds, if the government’s goal of confining the nationwide incidence of new cases over 7 days below 50 per 100,000 population were reached. I applied the latter figure in a sensitivity analysis. I arrived at the annual death toll of the pandemic (i.e., the number of deaths per pandemic wave) by halving the expected death toll of the first two pandemic waves in Germany.

The IFR was adjusted upwards to account for the long-term mortality of ICU survivors. The per capita gain in life years accounts for the percentage of the population that must be immune in order to reach the herd immunity threshold. Furthermore, given that the IFR is lower than the case fatality rate (CFR) in Germany, I adjusted the percentage of patients admitted to the ICU accordingly because a lower CFR also implies a lower percentage of cases admitted to the ICU (Gandjour 2020).

According to the United States Food and Drug Administration (FDA) (2020), the efficacy of the primary endpoint in a placebo-controlled efficacy trial should be at least 50%, to classify a widely deployed COVID-19 vaccine as effective, while ensuring safety. Hence, I took this estimate as the lower limit of the vaccine efficacy. As the FDA allows both SARS-CoV-2 infection and deaths associated with COVID-19 to be defined as primary endpoints, applying the 50% threshold to the life years gained as a measure of vaccine efficacy is still valid.

In both the Pfizer-Biontech and Moderna COVID-19 vaccines, efficacy in clinical trials in preventing the confirmed incidence of symptomatic COVID-19 was approximately 95%. While the efficacy of the vaccines against mortality needs to be confirmed, I took a 95% efficacy in preventing deaths as the upper limit of vaccine efficacy.

In the base case, I assumed that a vaccine campaign is able to overcome the vaccine hesitancy by using strategies such as simple, easy-to-understand language (Volpp 2020). Thus, the campaign was projected to achieve an uptake that is sufficient to yield the herd immunity. Based on equation 5 and a herd immunity threshold of 70% for natural infection (Kwok 2020), the threshold is approximately 73% for a vaccine efficacy of 95%. For a vaccine efficacy of 50% I assumed the same uptake.

In a sensitivity analysis, I considered a vaccine uptake of 50% based on a survey of November 2020 in the German population (Ärzteblatt 2020). If herd immunity is not reached, local outbreaks may follow, necessitating local shutdowns/lockdowns. The economic costs of the latter were already accounted for by the economic projections in the absence of another pandemic wave, because the projections assumed continuous spreading of infections and only a “gradual lifting of containment measures” (European Commission 2020).

Immunity was assumed to last between one (Galanti 2020) and ten years. The latter estimate is based on the immunity status of the survivors of SARS, caused by another coronavirus, who still carry certain important immune cells 17 years after their recovery (Le Bert 2020). However, the duration of immunity cancels out in the comparison between vaccination and no intervention, assuming that it is the same for both natural infection and immunization. For comparison between vaccination and mitigation, the GDP drops associated with annual pandemic waves under mitigation were discounted at an annual rate of 3%, based on the social rate of time preference derived from the Ramsey equation (Ramsey 1928). For health benefits of mitigation, I applied a 2% discount rate, reflecting a 1% expected growth rate of the consumption value of health in Germany (cf. John 2019).

### Willingness to pay (WTP)

The WTP for an additional life year (€101,493 per life year gained) based on the absolute rule was calculated by dividing the incremental costs of new, innovative cancer drugs (€39,751) by the incremental survival benefit (0.39 life years) (Gandjour 2020a). As the WTP estimate does not account for life extension costs, the latter were not considered in the pricing model either (variable *b* in equation 2).

WTP based on the proportional rule was derived from the ICER of the current mitigation strategy. Spending on the mitigation strategy may be seen as an appropriate reflection of its value given that the benefits and opportunity costs of the mitigation strategy have been intensely discussed in the public and the media.

## Results

Table 1 shows the input values and distributions used in the base case and sensitivity analysis. As shown in Table 2, a vaccine with 95% efficacy (with a price of zero) dominates the mitigation strategy strictly because it yields more life years at a lower cost. The value-based price based on an absolute rule (€1494) is thus determined by the comparison between vaccination and no intervention because the latter is the next most effective intervention. The value-based price refers to the complete vaccination course, i.e., it includes all doses. Furthermore, the proportional rule yields a price that is lower but within a close range.

**Table 1.** Input values and distributions used in the base case and sensitivity analysis.

| Input | Mean (range) | Reference |
| --- | --- | --- |
| *Epidemiological and clinical data* |  |  |
| Population size by age | see reference | Federal Office of Statistics 2020 |
| IFR in Germany | 0.0075 (0.005 – 0.01) | WHO 2020 |
| CFR in Germany |  | Robert Koch Institut 2020 |
| Total population | 0.021 |  |
| 0-9 years | 0.00015 |  |
| 10-19 years | 0.00029 |  |
| 20-29 years | 0.00013 |  |
| 30-39 years | 0.00028 |  |
| 40-49 years | 0.00099 |  |
| 50-59 years | 0.0034 |  |
| 60-69 years | 0.018 |  |
| 70-79 years | 0.073 |  |
| 80-89 years | 0.15 |  |
| 90+ years | 0.21 |  |
| Probability of ICU indication | 0.04 (0.04 – 0.08) | Robert Koch Institut 2020 |
| False-positive ICU admissions | 0.1 (0.1 – 0.2) | Abers 2014 |
| CFR in the ICU | 0.23 (0.22 – 0.24) | Robert Koch Institut 2020 |
| CFR one year post ICU discharge | 0.59 (0.47 – 0.73) | Damuth 2015 |
| Herd protection threshold | 0.70 (0.60 – 0.70) | Kwok 2020 |
| Immunity following one vaccination, years | 5 (1 – 10) | Galanti 2020, Le Bert 2020 |
| *Cost data* |  |  |
| GDP reduction per pandemic wave, % | 1.8 | European Commission 2020b |
| GDP reduction in 2020/21 without a second wave, % | 5.0 | European Commission 2020a |
| GDP drop attributable to shutdown versus voluntary restrictions, % | 100 (10 – 100) | Estimate |
| Construction and operation of vaccination centers, € | 1,000,000,000 | RND 2020 |
| Transport, storage, syringes, and needles, € | 231,000,000 | WDR 2020 |

CFR = case fatality rate, ICU = intensive care unit, IFR = infection fatality rate

**Table 2.**
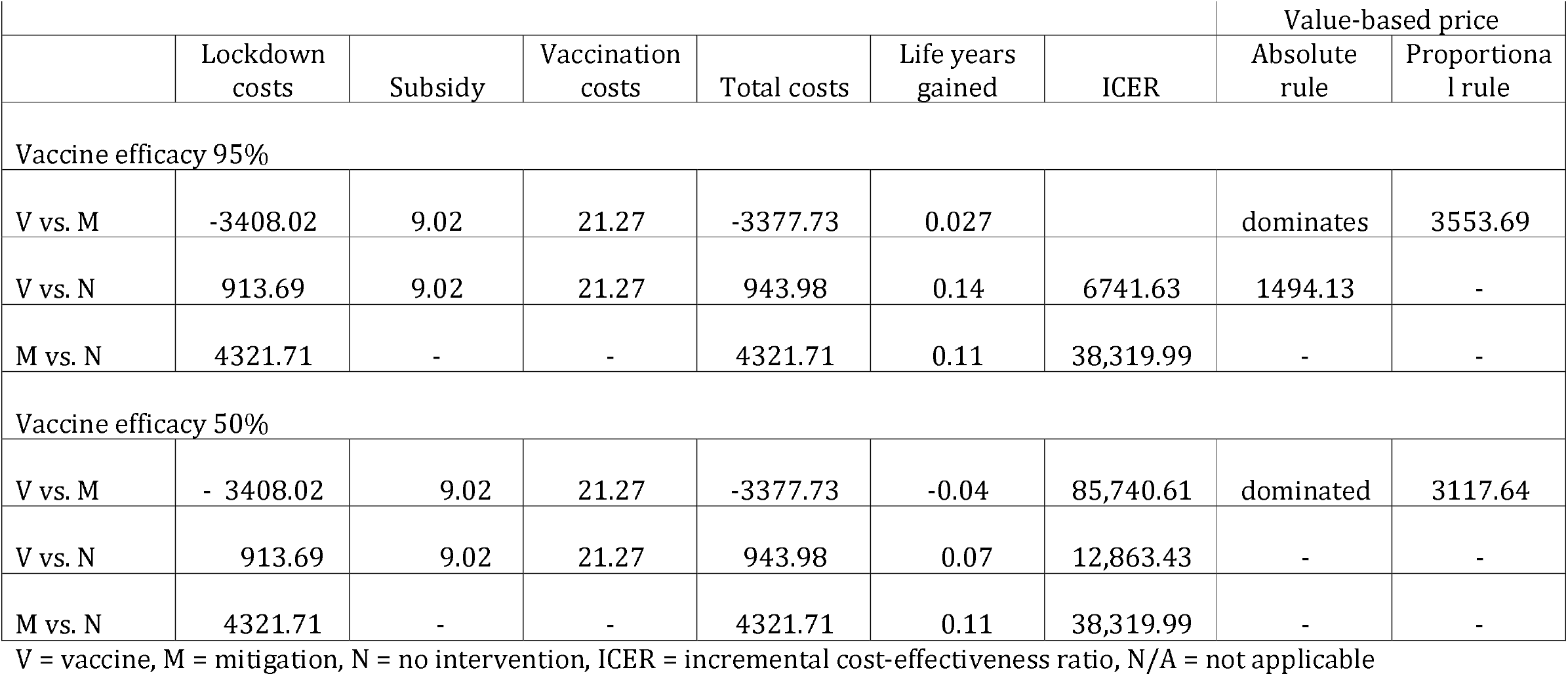
Value-based prices in the base case. All costs are in Euro. Costs and life years refer to one individual.

In contrast, a vaccine with 50% efficacy (with a price of zero) is less effective than the mitigation strategy. As the savings are not sufficiently large to pass the ICER threshold, vaccination is not cost-effective (the value-based price is zero). Nevertheless, if a vaccine with 50% efficacy is accompanied by partial lockdown measures in the long-term, based on the ICER of mitigation versus no intervention, the costs to achieve efficacy on par with long-term mitigation only will be 3.7% of GDP. The corresponding value-based price is $1868. A value-based price can also be determined based on the proportional rule because the loss of life years carries a lower weight due to the lower ICER threshold.

As shown in the sensitivity analysis (Figure 1), the range for the value-based price of a vaccine with a 95% efficacy, based on the absolute rule, lies between €678 and €3671. The major drivers of the price are the probability of an ICU admission and the GDP drop independent of the second wave (the former because of its considerable influence over the health outcomes under no intervention).

**Figure 1.**
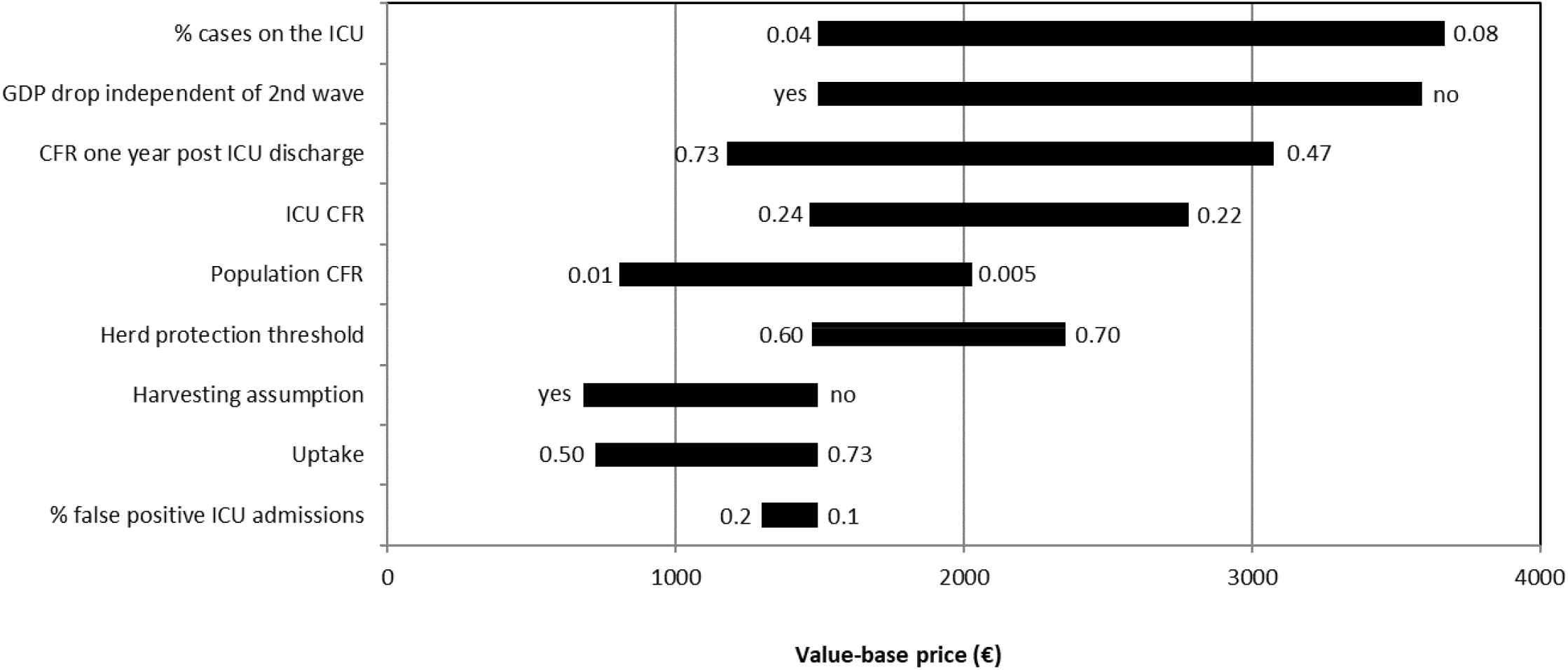
Tornado diagram demonstrating the results of the one-way sensitivity analysis. The variables are ordered by the impact on the value-based price of a COVID-19 vaccine based on an absolute rule (95% efficacy). The numbers indicate the upper and lower bounds.

## Discussion

This study yields a value-based price for a COVID-19 vaccine with 95% efficacy that is more than 50 times greater than the purchasing price even after incorporating the results of the sensitivity analysis (€1494 vs. €26 in the base case). In contrast, a vaccine with 50% efficacy has a negative value compared to a mitigation strategy. However, if the latter is not sustainable for economic, psychological, or other reasons, a vaccine with 50% efficacy is able to obtain a positive value compared to no intervention.

Towse and Firth (2020) argue that prices of COVID-19 vaccines need to be a value-based rather than cost-based to incentivise “a number of vaccines” and in particular “better vaccines” and “to avoid incentivising high cost low quality vaccines at the expense of lower cost, but better quality, ones.” In agreement, Neumann et al. (2020) make the case that not including the full value of a COVID-19 vaccine in its price encounters the risk of “having few effective products for the next pandemic”.

Towse and Firth (2020) suggest that the commitment to supply the vaccine on a not-for-profit basis must be time-limited because private investors expect a return. Hence, they propose suppling the vaccine on a not-for-profit basis only initially, but on “a normal commercial basis” in subsequent years. Based on this reasoning, the results of this study allow defining the potential for a price increase in the long-term. Otherwise, when VBP is not seen as realistic or desirable, the present results allow defining the portion of the price that reflects the contribution of the vaccine developers and the manufacturers to society, thus displaying their corporate social responsibility.

As a word of caution, given the time constraints and the rapid inflow of new information on the SARS-CoV-2 pandemic while conducting the study and writing this manuscript, which made it necessary to update the projections continuously, this decision-analytic study has several caveats. There are reasons why the study underestimates the health benefits and cost-effectiveness of a vaccine compared to the alternative strategies, and its value-based price. Some of these reasons were already captured in the sensitivity analysis and include a high probability of ICU admission. First, the study does not consider the deaths and loss of health-related quality of life associated with the shutdown and social distancing, e.g., due to depressive or anxiety disorders, suicides, unemployment, domestic violence, and fewer emergency and regular visits to physicians for unrelated medical conditions. Nevertheless, as mentioned in the Methods section, official data on excess mortality in Germany (Federal Office of Statistics 2020) show that the peak in excess mortality in the first half of April and the rising mortality in November both coincided with the surges in COVID-19 deaths, thus indicating that excess mortality was driven by COVID-19 and not by other causes. Second, unaffected individuals may experience a loss of personal freedom (Abele-Brehm 2020) and autonomy under lockdown. Third, under mitigation strategy elective procedures may need to be deferred if ICU capacity is expected to be insufficient. Forth, a vaccine may prevent COVID-19 infection with long-haul symptoms and save the direct (non-)medical costs and indirect costs associated with nonfatal COVID-19 cases. And fifth, under vaccination productivity gains resulting from reduced mortality were not included due to the age distribution of averted deaths (the median age is 83 years) and the difficulty of disentangling deaths in relevant age groups (e.g. in the age group 60–69 years).

Conversely, there are reasons to believe that the health benefits of mitigation may be underestimated and the value-based price of a vaccine may be overestimated. First, decreased economic activity can save lives, because it reduces air pollution, traffic accidents (Science Magazine 2020), and accidents on construction sites (Deaton 2020). Second, social distancing may reduce deaths due to non-COVID-19 flu. Third, a vaccine that is not 100% effective still allows COVID-19 infection with long-haul symptoms. Some of the biases listed in this and the previous paragraph may cancel each other out.

Furthermore, independent of questions around capturing the health benefits of the different strategies, it may be reasonable to subtract from the value-based price the portion of the vaccine’s value that is based on government-funded R&D costs (Neumann 2020). While the total subsidy by the German federal government for the three companies is known (€750 million), the total R&D spending incurred by the companies is not known.

Moreover, the extent to which the two mRNA-based vaccines reduce the likelihood of infection of SARS-CoV-2 upon exposure, i.e., efficacy of the vaccine on the susceptibility, is not yet known. A vaccine with 50% efficacy against confirmed disease, but lack of efficacy against the susceptibility of infection was projected to decrease COVID-related mortality by only 38%, based on a population uptake of 45% (Swan 2020).

Finally, the number of life years as an outcome measure may be criticized for lacking a consideration of health-related quality of life. Quality-adjusted life years (QALYs) are able to capture an additional health benefit resulting from the avoidance of non-fatal COVID-19 cases and the associated loss in quality of life. On the other hand, QALYs diminish the health benefits obtained from additional survival time by accounting for a quality-of-life decrement. As the QALY metric thus discriminates against the elderly and the disabled, it has been considered ethically controversial (Ubel 1999). For this and other reasons, QALYs have not been used so far in Germany for the purpose of reimbursing and pricing new, innovative medicines (cf. IQWiG 2020). As another counterpoint, the public debate on COVID-19 in Germany has mainly focused on mortality as an end-point and the number of life years lost by the elderly who die from COVID-19. In sum, there is not a straightforward answer to the question of which outcome measure best reflects the value of a successful vaccine. Life years gained may serve as a compromise between the use of unweighted lives saved and QALYs gained.

In terms of the transferability and relevance of the results and conclusions of this study to other countries, the usual caveats apply. The reasons for caution include between-country differences in clinical and epidemiological data, costs, and the willingness to pay for health benefits.

To summarize, this study demonstrates the applicability of VBP to a novel COVID-19 vaccine. In spite of the non-negligible uncertainties around the mean, the value-based price shows a considerable deviation from the cost-based price.

## Data Availability

All data are contained within the manuscript.

## Appendix

**Table A1.** Input data used for calculating the productivity loss due to an uncontrolled infection in the absence of a vaccine.

| Input | Mean (range) | Reference |
| --- | --- | --- |
| *Epidemiological* |  |  |
| IFR in Germany | 0.0075 | WHO 2020 |
| Percent of infections that are asymptomatic | 0.4 | CDC 2020 |
| Percent of diagnosed infections that are asymptomatic | 0.38 | Robert Koch Institut 2020 |
| Percent of diagnosed infections that are hospitalized | 0.07 | Robert Koch Institut 2020 |
| Quarantined contact persons per diagnosed case | 5 | Tagesschau 2020 |
| Number of newly diagnosed cases in August 2020 | 33,683 | Robert Koch Institut 2020 |
| *Cost data* |  |  |
| Hours worked per head and year in the population | 753.3 | OECD 2020 |
| Labor productivity per hour, € | 55.1 | OECD 2020 |

